# Histopathological Profile of Cervical Biopsies in Northern Malawi: A Retrospective Study

**DOI:** 10.1101/2020.12.28.20248949

**Authors:** Paul Uchizi Kaseka, Alfred Kayira, Chikondi Sharon Chimbatata, Master Chisale, Pocha Kamudumuli, Tsung-Shu Joseph Wu, BalwaniChingatichifwe Mbakaya, Frank Watson Sinyiza

## Abstract

**Objectives:** This study was carried out to determine the histopathological profile of cervical biopsies in a public tertiary hospital in Mzuzu, northern region of Malawi.

**Setting:** A public tertiary hospital in Mzuzu, northern region of Malawi

**Participants:** This was a retrospective study of all cervical biopsy specimen reports received in a public tertiary hospital in northern Malawi over a period of 5 years from July 2013-June 2018. Eleven reports which had missing demographic and clinical data or had inconclusive results were excluded. Demographic, clinical and histopathological data was obtained from original histology reports.

**Results:** A total of 500 cervical biopsy reports were reviewed during the study period. The mean age of the patients was 41.99±12.5. Age ranged from 15 to 80 years. Cervicitis accounted for 46.0% (n=162) of the total nonmalignant lesions seen, followed by cervical intraepithelial neoplasm (CIN), at 24.4% (n=86) and endocervical polyp, at 20.5% (n=72). Squamous cell carcinoma (SCC) accounted for 15.6% (n=78) of the total cervical biopsies studied and 85.7% of all total malignant lesions. All malignant tumours had HIV.

**Conclusion:** Our study shows that cervicitis and squamous cell carcinoma were most common among nonmalignant and malignant cervical biopsies respectively. Since the frequency of cervical cancer is high, there is need to have well detailed national policies to be put in place to increase detection of pre-invasive lesions in order to reduce the prevalence of cervical cancer.

**Strengths and limitation of this study Strengths:** This paper has shown

- The need for well detailed national policies to be put in place to increase detection of pre-invasive lesions, which in turn will decrease the frequency of cervical cancer in the country.
- The importance of intensifying cervical cancer screening programmes among women and provision of long term ART to the HIV infected which may offer an opportunity for appropriate interventions to reduce morbidity, mortality and reduce complications among these women.

**Limitations:**

- This study used available programme health facility data and histopathological reports on cervical cancer which has its own limitations, such as incompleteness and bias in the sense that information is obtained only from people who came to the facility and underwent biopsy, leaving out those that did not seek medical care and or were not biopsied and therefore cannot be generalized to the general population.
- The study is a single-hospital-based review and as such inadequate to draw conclusions, but it does shed some light on pathological pattern of cervical cancer in Malawi.

## BACKGROUND

Cervical cancer, after breast cancer, is the second most common cancer in women aged 15 to 44 years and it is the third leading cause of cancer in females worldwide(1). According to the International Agency for Research on Cancer (IARC) estimates, there are 570,000 new cases of cervical cancer annually, resulting into more than 311,000 deaths in 2018 globally (2). Most of the global burden lies in less developed countries, with sub-Saharan Africa (SSA) having the largest age-standardized incidence and mortality rates. Malawi has the highest cervical cancer incidence and mortality in the world with age-standardized rate (ASR) of 75.9 and 49.8 per 100,000 population respectively(3). World Health Organization (WHO) estimates suggest that every year there are at least 3,684 new cases of cervical cancer in Malawi and over 2,314 die from the disease(3).The exact number of cervical cancer morbidity and mortality among Malawian women, is not clear. This could partly be due to unrecorded or underreported cases because of the pathological based cancer registry not being maintained, and also lack of a national system of death certification(4). However, the 2010 National population-based cancer registry indicates that among females, cancer of the cervix was the commonest, accounting for 45.4% of all cases followed by Kaposis sarcoma (21.1%), cancer of the oesophagus (8.2%), breast cancer (4.6%) and non-Hodgkin lymphoma (4.1%)(5).

The standard method for diagnosis of cervical precancerous lesions is histopathological examination of tissue obtained through biopsy guided by colposcopy. When abnormalities are identified, cervical biopsy confirms the diagnosis of cancer(6).This study was carried out to determine the histopathological profile of cervical biopsies in a public tertiary hospital in Mzuzu, northern region of Malawi. The specific objectives were to determine the prevalence of both precancerous and cancerous cervical lesions; characterization of precancerous and cancerous lesions and risk factors of cervical cancer. Understanding the histological pattern of cervical cancer could guide development of focused preventive, care and treatment guidelines to address shortfalls in the care that is provided to cervical cancer patients in Malawi. Ultimately, this could contribute to the global target of 25% reduction of premature mortality from non communicable diseases (NCDs) by the year 2025.

## MATERIALS AND METHODS

### Design, setting and population

This was a retrospective study of all cervical biopsies reports received from Kamuzu Central Hospital/University of North Carolina (KCH/UNC) pathology laboratory in Lilongwe for Mzuzu Central Hospital (MCH), the only public tertiary hospital in the northern region of Malawi. This hospital does not have a functional pathology laboratory and relies on the KCH/UNC pathology laboratory for its services. The hospital is located in the northern part of Malawi catering for a population of about 2,289,780 million people (7) and serving 5 government District Hospitals, 3 Christian Health Association of Malawi (CHAM) hospitals and several private hospitals and clinics.

In this study a total of 500 cervical cancer pathology reports were analyzed over a period of 5 years (June 2013 to July 2018). Eleven reports which had missing demographic and clinical data or had inconclusive results were excluded. Data extracted included age, year, anatomic site, nature of specimen, clinical diagnosis, histopathological diagnosis, HIV status and whether the specimen was nonmalignant or malignant.

### Patient and public involvement

No patient involved

### Data analysis

Data were entered in Microsoft excel 2016, validated and cleaned before importing into Stata, version 13.0 (Stata Corp. LP, College Station, TX, United States of America) for analysis. Descriptive analyses were performed to summarize patients’ sociodemographic and clinical characteristics. Chi Square (or Fisher’s exact) test was used to look for significant associations between predictor and outcome variables at 95%significance level. A multiple simple logistic regression was used to quantify the association between predictor variables and outcome variables.

### Ethical clearance

This study was approved by National Health Science Research Committee (NHSRC) as part of the main study “Pathological profile of malignancies in northern Malawi-A retrospective study at Mzuzu Central Hospital” number 19/05/2316. Both the MCH Research and Publication Committee and the MCH Laboratory department consents were obtained for the study. The need for informed consent was exempted from the institutional review board due to the nature of the study, which involved a retrospective analysis of routinely collected data.

## RESULTS

Within the 5-year period of the study (July 2013 to June 2018), a total of 500 biopsy reports were received from KCH/ UNC Pathology laboratory. The mean age of patients included in this study was 41.99 ± 12.5. The age range was 15-80 years. Most of the cervical biopsies were from patients in the 31-40 age group (34.8% n=174). Twelve percent of the cervical biopsies were from HIV positive patients and 51% of the results had unknown HIV result (Table 1).

**Table 1.**
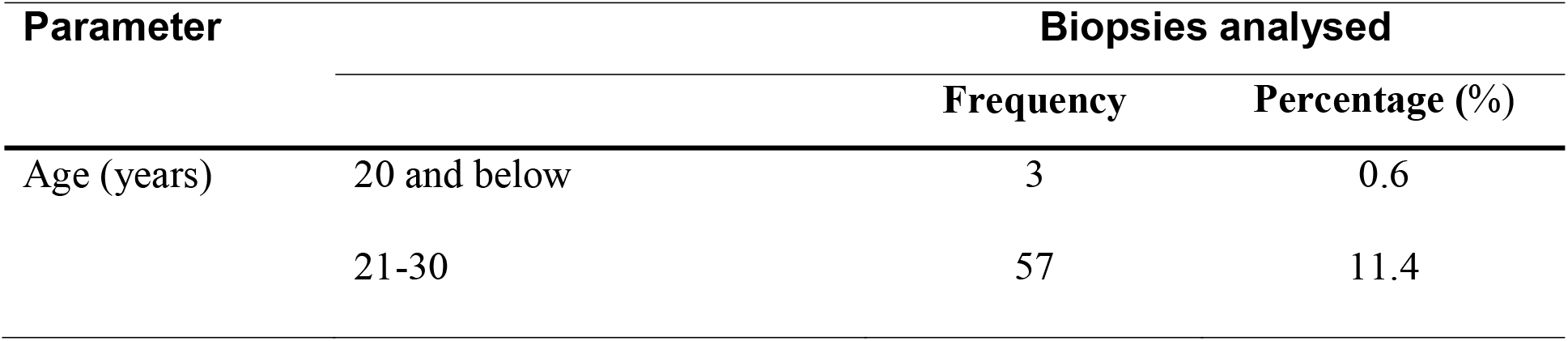
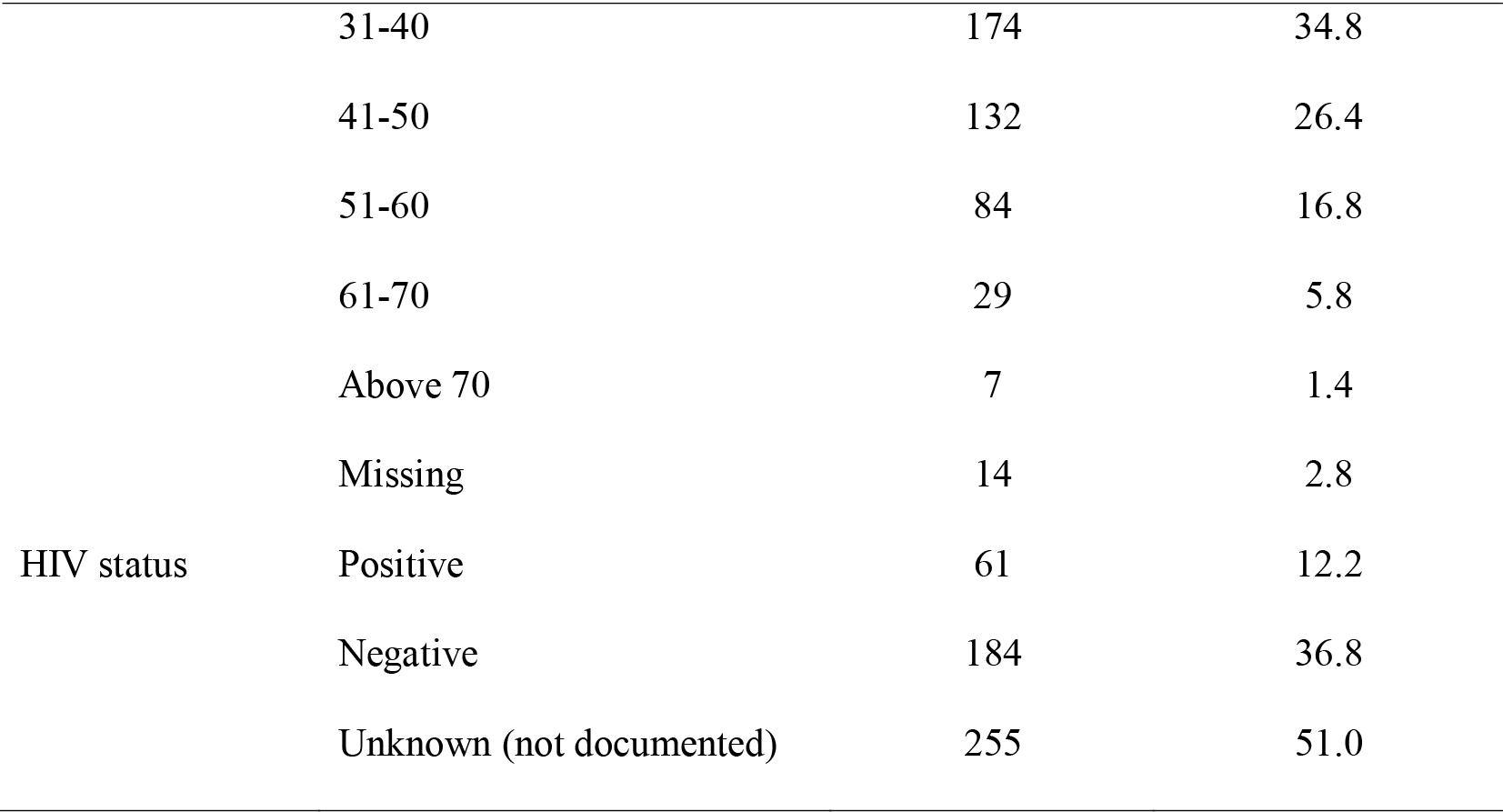
Demographic profile of cervical biopsies.

Of all the cervical biopsies studied, 91 (18.2%) were malignant. Squamous cell carcinoma (SCC) accounted for 85.7% of all malignant lesions (Table 2). Cervicitis accounted for 46.0% (n=162) of the total nonmalignant lesions seen. Ten percent of cervicitis cases and 30.2% of CIN cases had HPV respectively (Table 2).

**Table 2.** Histopathological diagnosis of cervical biopsies.

| Parameter | Frequency | Within group percentage(%) | Percentage(%)total |
| --- | --- | --- | --- |
| **Normal cervical tissue** | **46** |  | **9.2** |
| **Nonmalignant lesions** | **352** |  | **70.4** |
| Cervicitis* | 162 | 46.0 | 32.4 |
| Cervical intraepithelial neoplasm (CIN)* | 86 | 24.4 | 17.2 |
| Endocervical polyp | 72 | 20.5 | 14.4 |
| Carcinoma in situ | 4 | 1.1 | 0.8 |
| Narborthian cyst | 4 | 1.1 | 0.8 |
| Schistosomiasis | 5 | 1.4 | 1.0 |
| Condloma (warts) | 11 | 3.1 | 2.2 |
| Others | 8 | 2.3 | 1.6 |
| **Malignant** | **91** |  | **18.2** |
| Squamous cell carcinoma | 78 | 85.7 | 15.6 |
| Adenocarcinoma | 8 | 8.8 | 1.6 |
| Poorly differentiated | 4 | 4.4 | 0.8 |
| Carcino-sarcoma | 1 | 1.1 | 0.2 |
| **Inconclusive result** | **11** |  | **2.2** |
| **Total** | **500** |  | **100** |

\*Cervicitis with HPV= 10% (16/162)
\*CIN with HPV= 30.2% (26/86)
\*CIN 1 = 34.9% (30/86), CIN 2 = 22.1% (19/86), CIN 3 = 43.0% (37/86)

The chances of developing cervical cancer increased with advancing age, with the risk becoming more prominent after age 60 years. Women aged 61-70 years were 6.2 times (OR = 6.2, 95% CI:2-1 - 18.4) more likely to develop cervical cancer than those in the 21-30 years’ age category. The odds of getting cancer were higher in women living with HIV. Women with HIV were more than twice (OR = 2.3, 95% CI: 1.1 - 4.9) as likely to have cervical cancer as those who were HIV negative (Table 3).

**Table 3:**
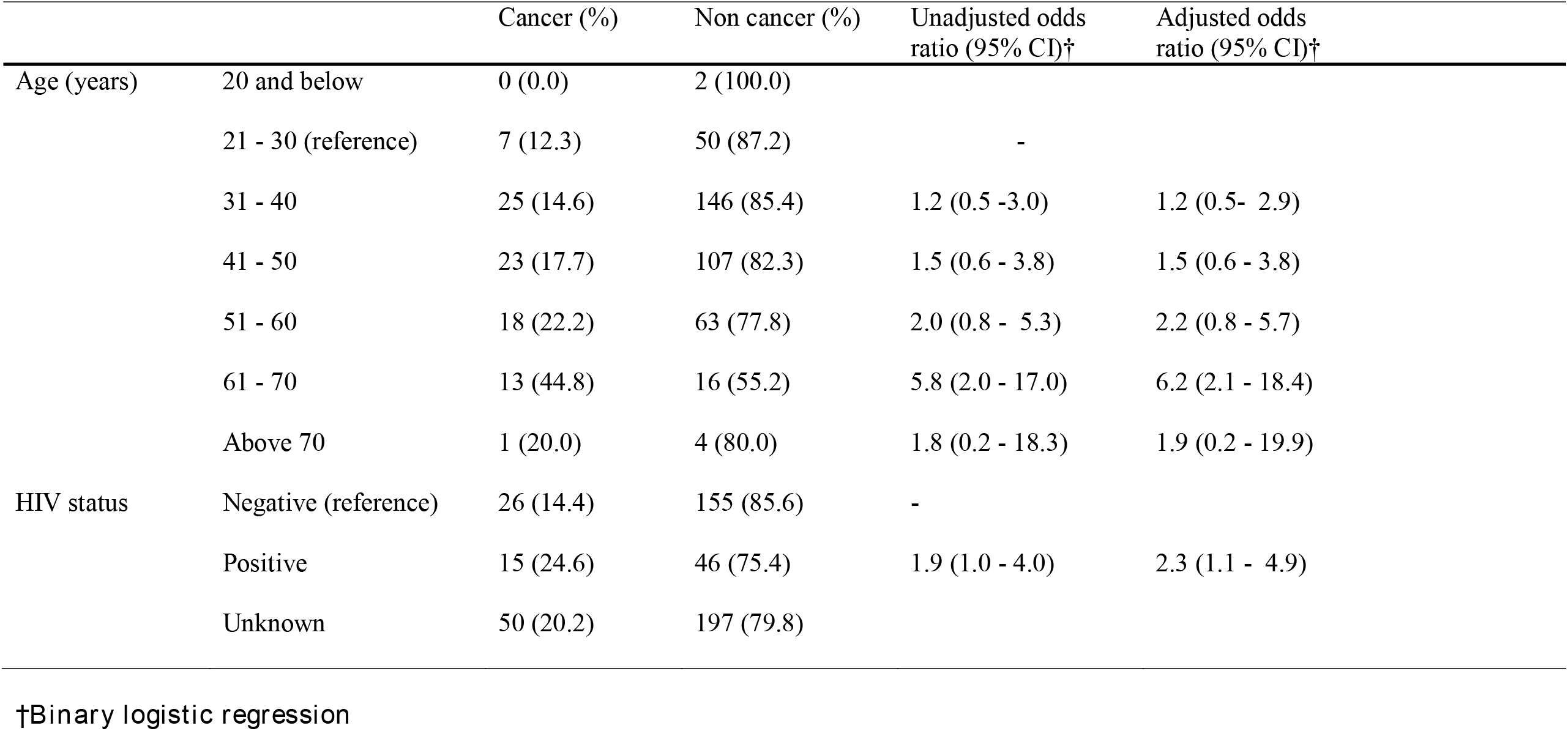
Association between cancer, age and HIV status.

## DISCUSSION

To the best of our knowledge, this is the first study conducted to determine the histopathological profile of cervical biopsies in a public tertiary hospital in Mzuzu, northern region of Malawi. This study found more benign conditions (70.4% n=352) than neoplastic malignant conditions (18.2% n=91). This is in contrast with other studies undertaken in developing countries which reported more neoplastic malignant conditions than benign conditions(8,9).

Our study found that cervicitis, an inflammatory disease comprising acute cervicitis and chronic non-specific cervicitis was the most common nonmalignant condition. It accounted for 46% of all the nonmalignant tumours and 32.4% of all cervical biopsies received in this study. This was consistent to a study conducted in Nigeria whose cervical biopsies included in their study was 37.5%(8). Most cases of cervicitis are often due to non-specific causes or infective agents(9). Cervicitis has been highly reported in previously studies in other countries(12,13).

In this study CIN, a premalignant lesion ranked second in nonmalignant tumours accounting for 24% of all nonmalignant tumours and 17.2% of all cervical biopsies with the prevalence of CIN I, CIN II, CIN III of 34.9%, 22.1% and 43% respectively. This finding is comparable to a study conducted in Nigeria which reported CIN constituting 15% of all cervical biopsies reviewed(10). In that study however, there was a reduction in the prevalence of CIN from low grade CIN to high grade CIN(10). In our study, the high grade CIN (CIN III) was the most common. This finding indicates that invasive cervical cancer progresses from advanced stages of precancerous lesions. This shows that there is need to create community awareness and strengthen early cervical cancer screening for Malawi to have better outcomes.

In the current study 10% (n=16) of all the cervicitis cases and 30.2% (n=26) of all the CIN biopsies had HPV. This is comparable to a study conducted in United States of America (USA) which reported the overall HPV prevalence of 26.8% among US females aged 14 to 59 years (n=1921)(11). This finding suggests that there is high rate of HPV infections, the causative agent of CIN in the younger age group(10). Unfortunately, the data on population-based, age specific prevalence of HPV are not available in Malawi. However, WHO estimates that the overall HPV prevalence in Malawi is about 34%(12).

The rate of malignant lesions (tumours) in our study (18.2%) is comparable to 16.2% and 12.2% in studies conducted in Benin City(13), and in Enugu, in Nigeria(14) respectively. Studies done in South Africa, Saudi Arabia, India and United States, with malignant lesions at 2.42%, 4.95%, 5.5% and 5.0% respectively, are all at variance with the current study which explains that early cervical cancer screening helps to reduce the prevalence of cervical cancer(15–18).

Among the malignant tumours, SCC was the most common histological type of cervical cancer in our study. SCC accounted for 85.7% of malignant lesions cervical cancer and was also the most common diagnosis, at 15.6% of all cervical biopsies in this study. This is consistent with findings from other studies conducted in Nepal and in Pakistan which reported 84.2%, 92.56%, and 73.5% respectively for squamous cell carcinoma out of all malignant lesions reviewed(19–21). As Faduyile et al. observed the high rate of SCC in Malawi and Africa could reflect the low uptake of VIA and Pap smear test which are capable of identifying dysplastic conditions before transformation to malignancy(11). This shows that there is need for Malawi to have well organized cervical cancer screening and Pap smear test to reduce the prevalence of SCC(2).

In our study, adenocarcinorma and poorly differentiated carcinorma were 8.8% and 4.4% respectively of the total malignant diagnosis. The prevalence of adenocarcinoma and poorly differentiated carcinoma observed in this study is in contrast to other studies done in Nigeria where one study found 5.8% and 2.0% of the total malignant lesions diagnosis and another found 6.0% and 1.0% respectively(10,22). The high prevalence found in this study confirms a previous study findings by Chanza et al.(23). Chanza et al. reported that Malawian women delay to seek medical attention due to limited knowledge on symptoms and signs, limited financial resources, limited accessibility and unavailability of cancer screening facilities hence late diagnosis. These results show that early cervical cancer screening, increased awareness, better health care facilities, accessibility, improved histopathological confirmatory diagnosis and early treatment by surgeons, may reduce the cervical cancer burden in Malawi.

In the current study it has been observed that the diagnosis of cervical cancer was significantly associated with the age of the patients. The chances of developing cervical cancer increased with advancing age, with the risk becoming more prominent after age 60 years. Women aged 61-70 years were 6.2 times (OR = 6.2, 95% CI: 2.1 - 18.4) more likely to develop cervical cancer than those in the 21-30 years age category (Table 3). It is important to note that high quality screening programs are important to prevent cervical cancer among unvaccinated older women(24). None of the women in our study had received any HPV vaccine in their lifetime as the first round of the mass HPV vaccine in this country was administered in January 2019 mainly in schools, targeting 9-13 year old girls who had not yet become sexually active and the second round was given in January 2020. In Malawi, the integration of HPV vaccine programs with adequate screening programs in older women (aged 30-49 years) has the potential to reduce the burden of cervical cancer.

Malawi’s HIV prevalence is one of the highest in the world, with 10.6% of adult population (aged 15-64) living with HIV (25). With such a high HIV prevalence in Malawi, there is an increased risk of AIDS-defining cancers including cervical cancer. In this study the probability of getting cancer were higher in women living with HIV. Women with HIV were more than twice (OR = 2.3, 95% CI: 1.1 - 4.9)as likely to have cervical cancer as those who were HIV negative (Table 3).This could be attributed to the reason that HIV-infected women are more likely than HIV-uninfected women to have incident and persistent HPV cervical infections(26). A twelve monthly cervical cancer screening, increased availability of dolutegravir (DTG) based antiretroviral treatment (ART) to HIV infected women now being provided in the country, routine viral load checks (at 6 months, 12 months and every 12 months since initiation) and timely switch to second line or third line regimens to ensure viral load suppression will eventually have an impact on benign lesions progressing to invasive cervical cancer(27). However, in this study almost half (52.74%, n=48) of all the women diagnosed with cervical cancer had an unknown HIV status. This calls for scaling up of HIV testing in cancer screening settings for early diagnosis and ART referral and further research is warranted on barriers among HIV-infected women to seeking cancer screening services despite already being in the health care system.

## LIMITATIONS

This study used available programme health facility data and histopathological reports on cervical cancer. The use of health facility data has its own limitations, such as incompleteness and bias in the sense that information is obtained only from people who came to the facility and underwent biopsy, leaving out those that did not seek medical care and or were not biopsied and therefore cannot be generalized to the general population.

The other limitation of this study is that it is a single-hospital-based review and as such inadequate to draw conclusions, but it does shed some light on pathological pattern of cervical cancer in Malawi.

Finally, this is a retrospective study so we could not be able to extract details for example in cases where tumours were diagnosed by screening or symptoms, presence or absence of the patient co-morbidities. Nevertheless, the comprehensive histopathological pattern of cervical cancer demonstrated by this study provides evidence that could be used to inform policies, strategies and intervention for prevention of cancer in Malawi.

## CONCLUSION

The SCC was the commonest malignant condition and cervicitis and CIN were the most common non malignant conditions in all the women studied. Since the frequency of cervical cancer is high, there is need for well detailed national policies to be put in place to increase detection of pre-invasive lesions, which in turn will decrease the frequency of cervical cancer in the country. The presence of chronic non-specific cervicitis in women of reproductive age is infective in origin with its attending sequelae. Intensifying screening programmes among women and provision of long term ART to the HIV infected may offer an opportunity for appropriate interventions to reduce morbidity, mortality and reduce complications among these women.

## Data Availability

All data relevant to the study are included in the article and
Data are available upon reasonable request from the corresponding author using the following contacts
ORCID: 0000-0002-6651-8000

## Declarations

### i. Ethical approval and consent to participant

This study was approved by National Health Science Research Committee as part of the main study “Pathological profile of malignancies in northern Malawi-A retrospective study at Mzuzu Central Hospital” number 19/05/2316. Both the Mzuzu Central Hospital Research and Publication Committee and the Mzuzu Central Hospital Laboratory department consents were obtained for the study. The need for informed consent was exempted from institutional review board due to the nature of the study, which involved a retrospective analysis of routinely collected data.

### ii. Consent for publication

Not applicable

### iii. Availability of data and materials

The datasets used and/or analyzed during the current study are available from the corresponding author on reasonable request.

### vi. Authors contributions

PUK and FWS conceived and designed the study. AK, CC, PK, JU and BCM contributed to development of the study protocol and supervised data collection and entry. AK analyzed the data and PUK drafted the manuscript. All authors read and approved the final manuscript.

## vii. Acknowledgements

The authors are sincerely grateful to all the data collectors and the Laboratory manager for providing the data. The Pingtung Christian Hospital through Luke International Norway (LIN) is also appreciated for financial support

